# Challenges in Building and Maintaining Rare Disease Patient Registries: Results of a Questionnaire Survey in Japan at 2012

**DOI:** 10.1101/19004770

**Authors:** Mizuki Morita, Soichi Ogishima

**Affiliations:** Center for Knowledge Structuring, The University of Tokyo < >; Department of Bioclinical Informatics, Tohoku Medical Megabank Organization, Tohoku University < >

**Keywords:** Needs, Rare diseases, Patient registries, Survey, Sustainability

## Abstract

**Background:** Research infrastructure such as patient registries and biobanks is expected to play important roles by aggregating information and biospecimens to promote research and development for rare diseases. However, both building and maintaining them can be costly. This paper presents results of a survey of patient registries for rare diseases in Japan conducted at the end of 2012, with emphasis on clarifying costs and efforts related to building and maintaining them.

**Results:** Of 31 patient registries in Japan found by searching a database of research grant reports and by searching the internet, 11 returned valid responses to this survey. Results show that labor and IT system costs are major expenses for developing and maintaining patient registries. Half of the respondent patient registries had no prospect of securing a budget to maintain them. Responders required the following support for patient registries: financial support, motivation of registrants (medical doctors or patients), and improved communication with and visibility to potential data users. These results resemble those reported from another survey conducted almost simultaneously in Europe (EPIRARE survey).

**Conclusions:** Survey results imply that costs and efforts to build and maintain patient registries for many rare diseases make them unrealistic. Some alternative strategy is necessary to reduce burdens, such as offering a platform that supplies IT infrastructure and basic secretariat functions that can be used commonly among many patient registries.

## Introduction

Grasping an overview of a single rare disease is difficult because of the lack of sufficient information related to it. That difficulty inhibits proper diagnosis and appropriate treatment, while impeding the progress of research related to that disease. Patient registries and biobanks are expected to alleviate some difficulties by gathering dispersed information and biospecimens and thereby drive rare disease research [Lopez et al. 2016].

Patient registries are usually intended for long-term operation. However, understanding how much effort is necessary to develop and maintain them is difficult. Several good reviews and recommendations written by experts in the field of rare diseases and patient registries have been published on this topic to date [Richesson & Vehik 2010; RDTF 2011; EUCERD 2013]. They include points that should be considered in establishing patient registries. New developers of rare disease patient registries can refer to this literature, but it does not include precise information that developers often demand. Sharing experiences of building and maintaining existing patient registries would be useful not only for future patient registries but also for existing patient registries. In Europe, the European Platform for Rare Diseases Registries (EPIRARE) project, 2011–2013 [EPIRARE] administered a large-scale online survey targeted mainly at patient registries in Europe (the EPIRARE Survey [EPIRARE Survey]) and published the results in 2013 [EPIRARE 2013]. The EPIRARE Survey asked about major needs of patient registries, ways to address various (methodological, technical, and regulatory) issues, and ways to find resources (technological tools, specific expert advice, model documents, and quality control systems) to develop and sustain patient registries. Reportedly, rare disease patient registries in Europe reported that their major needs include 1) gathering financial support, 2) motivating participants (data providers), 3) assessing data quality, and 4) improving communication and visibility of the obtained results.

This paper reports the results of a survey of rare disease patient registries in Japan conducted in 2012, which specifically addressed highlighting challenges that researchers face in constructing and maintaining patient registries of rare diseases, aiming at providing basic information that would be referred by existing and future patient registries.

## Methods

### Respondent selection

A list of potential survey participants was developed from a structured search of published sources through Google and the MHLW Grants System [MHLW Grants System], a database of research projects granted by the Ministry of Health, Labour and Welfare (MHLW) in Japan after 1997. When this survey was conducted, system users could search only the titles and abstracts of research reports submitted to the MHLW. Database searches using Japanese equivalent terms of “patient registry” were applied to the MHLW Grants System and Google. For all search results from the MHLW Grants System, we read abstracts of research reports to confirm the description that a patient registry(s) was constructed in the study. This process took place during August–October 2012.

From this search procedure, 31 potential patient registries in Japan were identified (26 from the MHLW Grants System and 5 from Google). Because only insufficient information can be captured from the title and abstract of research reports recorded in the MHLW Grants System, in most cases we were unable to ascertain the subject diseases of the potential patient registries. Therefore, the candidate list from the MHLW Grants System might include patient registries targeting non-rare diseases.

### Questionnaire management

A postal questionnaire with a request letter and a self-addressed envelope was sent to all of these potential patient registries in November 2012. Several days after the questionnaire sent, a phone call was made to ensure the arrival of the questionnaire. One month after the questionnaire was sent out, a reminder was sent to each addressee. To shed light on issues and challenges in building and sustaining rare disease patient registries in Japan, the following categories of questions were included: general characteristics of registries, registration method of patients, operating organization of registries, the budget for building, and sustaining registries.

## Results

### Respondent characteristics

Of the 31 subjects surveyed, 13 returned their questionnaires. Of those respondents, 11 completed the questionnaires. Two reported that registries had not been constructed. Of the 11 patient registries, 6 were for rare diseases, and 5 were for common diseases. Among the six respondent patient registries for rare diseases, three registries were established in 2009. Also, one registry was established in each of 2007, 2010, and 2011. Half of the six registries’ respondents reported that they have been cooperating with foreign registries or international networks.

The objectives of the registries are presented in Table 1. Epidemiological surveys were included in the objectives of all registries. The objectives related to diagnosis and treatment of the target diseases were considered by more than half of the registries. One objective related to drug development, “clinical trial of drug or medical device,” was considered by more than half of the registries, although “post-marketing surveillance of drug or medical device” and “market survey of drug or medical device” were considered by only one registry. The objectives related to a patient’s daily life, “evaluation of the quality of medical care,” “evaluation of economics of medical care,” and “survey of the living condition of patients,” were considered by only one or no registry. Registries 1 and 6 narrowed the objectives, although registries 3, 4, and 5 covered a wide ranges of objectives.

**Table 1.** Objectives for establishing patient registries for rare diseases

|  | Patient registry |  |  |  |  |  | Count |
| --- | --- | --- | --- | --- | --- | --- | --- |
|  | #1 | #2 | #3 | #4 | #5 | #6 |  |
| Epidemiology |  |  |  |  |  |  |  |
| 1. Prevalence and morbidity | + |  |  | + | + |  | 3 |
| 2. Symptoms, severity and complications | + | + | + | + | + |  | 5 |
| 3. Age of onset, gender difference and frequent site of onset | + | + | + | + | + |  | 5 |
| 4. Natural history | + |  | + | + | + | + | 5 |
| Diagnosis and treatment |  |  |  |  |  |  |  |
| 5. Establishment or evaluation of diagnostic criteria |  | + |  | + |  |  | 2 |
| 6. Establishment or evaluation of clinical guidelines |  | + |  | + | + |  | 3 |
| 7. Evaluation of treatment |  | + | + | + | + |  | 4 |
| Drug development |  |  |  |  |  |  |  |
| 8. Clinical trial of drugs or medical devices |  |  | + | + | + | + | 4 |
| 9. Post-marketing surveillance of drugs or medical devices |  |  |  |  | + |  | 1 |
| 10. Market survey of drug or medical device |  |  |  |  | + |  | 1 |
| Others |  |  |  |  |  |  |  |
| 11. Evaluation of medical care quality |  |  |  | + |  |  | 1 |
| 12. Evaluation of medical care economics |  |  |  |  |  |  | 0 |
| 13. Survey of patient living conditions |  |  | + |  |  |  | 1 |
| 14. Others |  |  | + |  |  |  | 1 |

### Registration method of patients

For three out of six respondent registries, medical doctors register in the registry under a patient’s consent. For the other three registries, patients or their representatives register directly. Registration is available only by postal mail in four respondent registries, only through the internet in one registry, and by both ways in one registry.

### Operating organization of registries

Offices of the patient registries were located in medical or research institutions for four registries, in a patients’ association in for one registry, and in a private company for one registry. The numbers of total staff in registry offices were 1, 4, 8, and 10 for four registries, and 6 for two registries. The numbers of full-time staff were 0 for three registries, and 1, 2, or 4 for each other registry.

### Budget for building and sustaining registries

The total budgets for establishing and maintaining each patient registry are presented in Table 2. For both establishment costs and maintenance cost of the preceding year, 3 of 6 respondent registries spent between 1 million and 5 million JPY (almost 10,000 and 50,000 EUR), 2 registries spent between 5 million and 10 million JPY (almost 50,000 and 100,000 EUR), and 1 registry spent between 10 million and 50 million JPY (almost 100,000 and 5,000,000 EUR). In the EPIRARE survey, the number of registries with yearly budget up to 100,000 EUR was 74 (36.8% of total of 201 respondents) and that with higher than 100,000 EUR was 36 (17.9%) [EPIRARE Survey]. JPY stands for Japanese Yen and EUR stands for Euro. Maintenance costs were of the prior year. The yearly average exchange rate in 2012 between JPY and EUR was 1 EUR to 102.13 JPY, calculated as the average between the Yearly Average Telegraphic Transfer Selling rate (TTS) and the Yearly Average Telegraphic Transfer Buying rate (TTB) obtained from the final official quotation by the Bank of Tokyo-Mitsubishi UFJ (BTMU) [MURC].

**Table 2.**
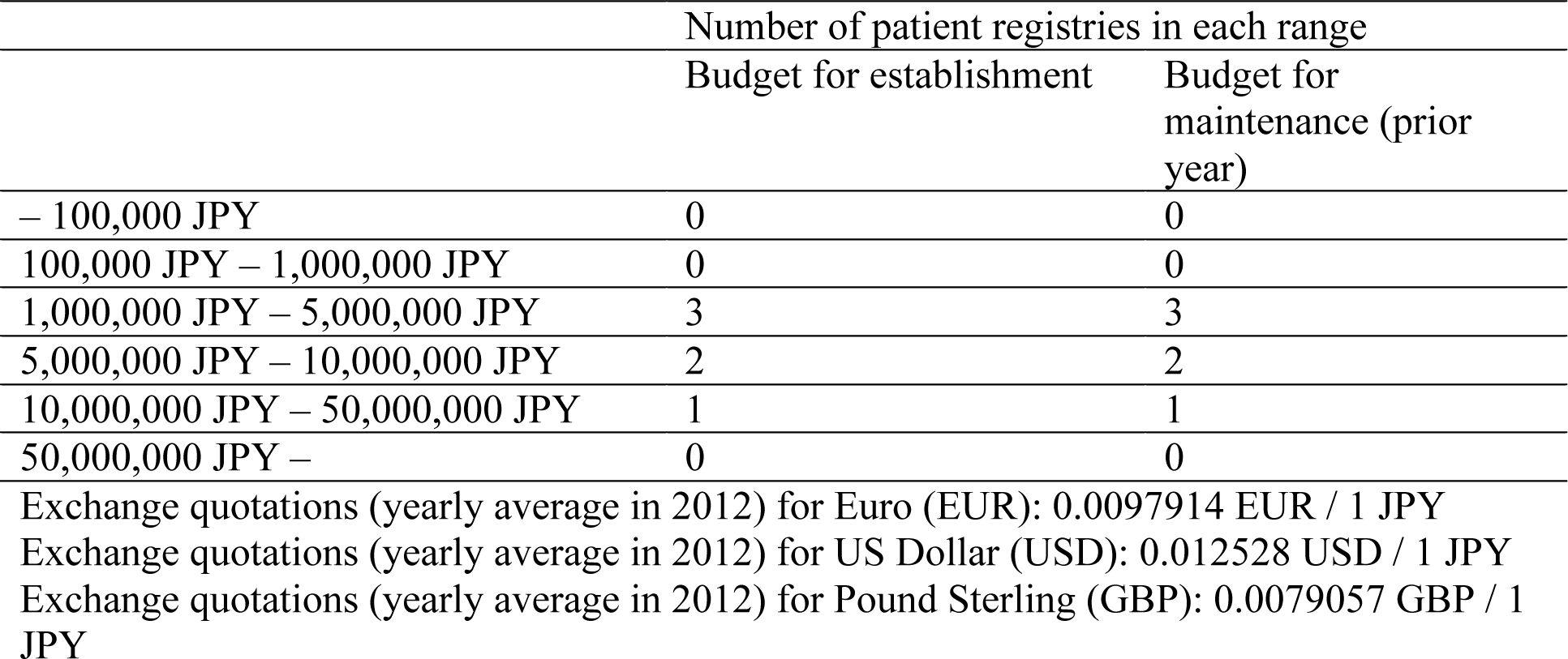
Budgets actually used for establishing and maintaining each patient registry in Japanese Yen (JPY)

The Top 3 funding and expense items for establishing each patient registry are presented in Table 3a and Table 4a. Four of the six registries listed the public research grant from Ministry of Health, Labour and Welfare (MHLW) as the top funding items in establishment of the patient registry. This result reflects that many medical studies are funded by this grant, and also that the MHLW Grants System and Google were used to search patient registries in this study. The other two registries listed financial assistance from the administering organization at the top 1. One registry listed donations; another registry listed research and funding by the university as the top 2. For expense items in establishment of the patient registry, the labor costs and information technology (IT) system construction costs were included in the top 2 in most registries. All registries listed one of these two items at the top 1. All except for one registry listed the remainder of them in the top 2. Many registries listed equipment and supplies expenses among the top 3, but no registry listed office expenses and energy (light, fuel and water) expenses within the top 3.

**Table 3.**
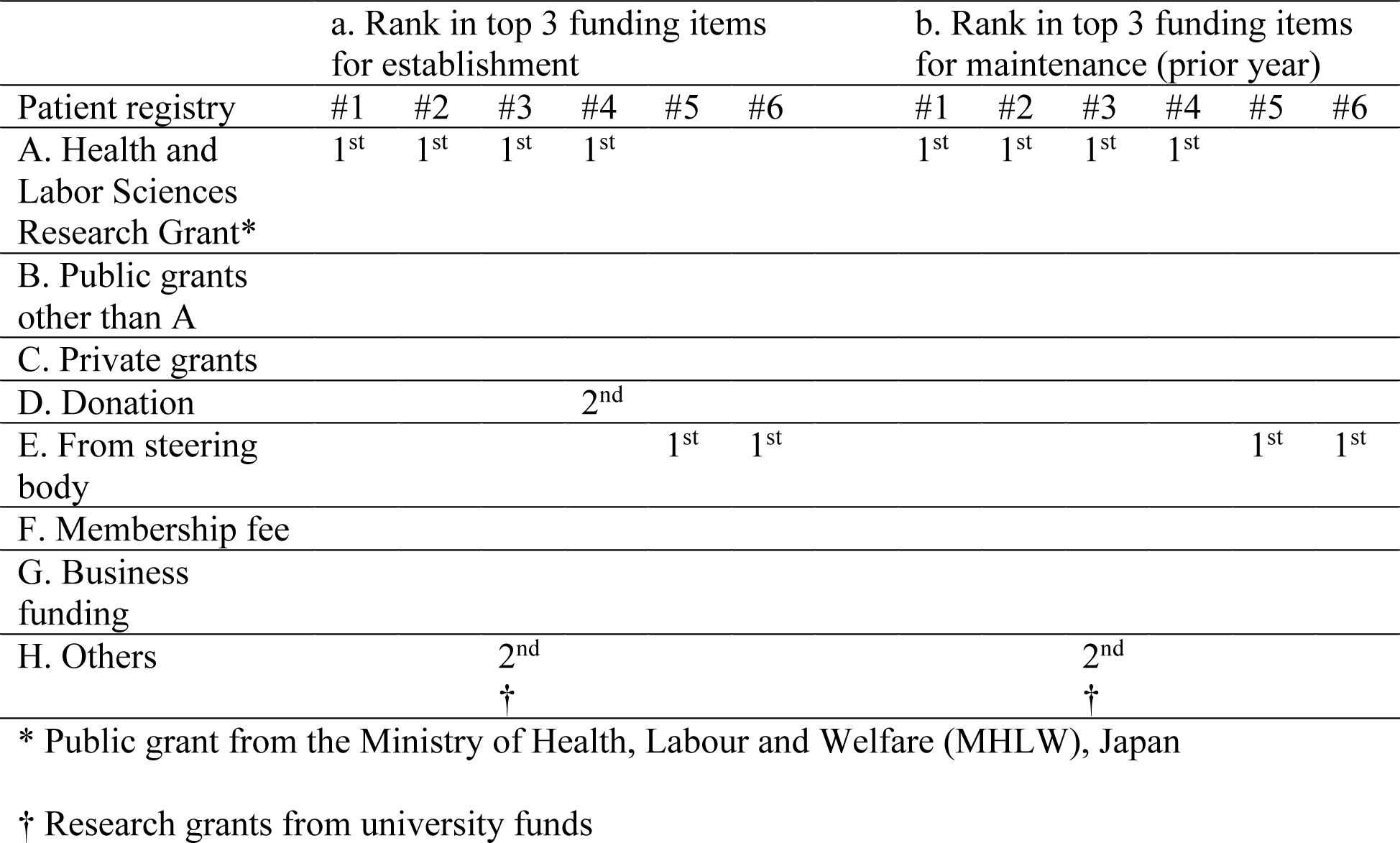
Top 3 funding items for establishing and maintaining each patient registry

|  | a. Rank in top 3 funding items<br>for establishment |  |  |  |  |  | b. Rank in top 3 funding items<br>for maintenance (prior year) |  |  |  |  |  |
| --- | --- | --- | --- | --- | --- | --- | --- | --- | --- | --- | --- | --- |
| Patient registry | #1 | #2 | #3 | #4 | #5 | #6 | #1 | #2 | #3 | #4 | #5 | #6 |
| A. Health and<br>Labor Sciences<br>Research Grant* | 1<sup>st</sup> | 1<sup>st</sup> | 1<sup>st</sup> | 1<sup>st</sup> |  |  | 1<sup>st</sup> | 1<sup>st</sup> | 1<sup>st</sup> | 1<sup>st</sup> |  |  |
| B. Public grants<br>other than A |  |  |  |  |  |  |  |  |  |  |  |  |
| C. Private grants |  |  |  |  |  |  |  |  |  |  |  |  |
| D. Donation |  |  |  | 2<sup>nd</sup> |  |  |  |  |  |  |  |  |
| E. From steering<br>body |  |  |  |  | 1<sup>st</sup> | 1<sup>st</sup> |  |  |  |  | 1<sup>st</sup> | 1<sup>st</sup> |
| F. Membership fee |  |  |  |  |  |  |  |  |  |  |  |  |
| G. Business<br>funding |  |  |  |  |  |  |  |  |  |  |  |  |
| H. Others |  |  | 2<sup>nd</sup> |  |  |  |  |  | 2<sup>nd</sup> |  |  |  |
|  |  |  | † |  |  |  |  |  | † |  |  |  |

\* Public grant from the Ministry of Health, Labour and Welfare (MHLW), Japan
† Research grants from university funds

**Table 4.** Top 3 expense items for establishing and maintaining each patient registry

|  | a. Rank in top 3 expense items<br>for establishment |  |  |  |  |  | b. Rank in top 3 expense items<br>for maintenance (prior year) |  |  |  |  |  |
| --- | --- | --- | --- | --- | --- | --- | --- | --- | --- | --- | --- | --- |
| Patient registry | #1 | #2 | #3 | #4 | #5 | #6 | #1 | #2 | #3 | #4 | #5 | #6 |
| A. Personnel | 1<sup>st</sup> | 1<sup>st</sup> | 2<sup>nd</sup> | 2<sup>nd</sup> | 1<sup>st</sup> | 1<sup>st</sup> | 1<sup>st</sup> | 1<sup>st</sup> |  | 2<sup>nd</sup> | 1<sup>st</sup> | 1<sup>st</sup> |
| B. Light, fuel and<br>water |  |  |  |  |  |  |  |  | 1<sup>st</sup> |  |  | 2<sup>nd</sup> |
| C. Equipment and<br>supplies | 2<sup>nd</sup> |  | 3<sup>rd</sup> | 3<sup>rd</sup> | 3<sup>rd</sup> | 3<sup>rd</sup> | 2<sup>nd</sup> |  |  | 3<sup>rd</sup> | 3<sup>rd</sup> |  |
| D. Office expenses |  |  |  |  |  |  |  |  |  |  |  |  |
| E. IT system | 3<sup>rd</sup> | 2<sup>nd</sup> | 1<sup>st</sup> | 1<sup>st</sup> | 2<sup>nd</sup> | 2<sup>nd</sup> | 3<sup>rd</sup> | 2<sup>nd</sup> | 2<sup>nd</sup> | 1<sup>st</sup> | 2<sup>nd</sup> |  |
| F. Others |  |  |  |  |  |  |  |  | 3<sup>rd</sup> |  |  | 3<sup>rd</sup> |
|  |  |  |  |  |  |  |  |  |  |  |  | * |

\* Communications expenses

The top 3 funding and expense items for maintaining each patient registry were presented in Table 3b and Table 4b. The research grant from MHLW was also the major funding item in maintaining the patient registries as well as in establishing them. However, because this grant funding expires typically in several years, it can not be regarded as a permanent funding source. Therefore, major funding items for maintaining patient registries after years can differ according to patient registries. Most patient registries included labor costs and IT system maintenance costs among the top 2 expenses for maintaining patient registries. One patient registry did not include labor costs. Two patient registries did not include IT system maintenance costs among the top 3 expenses.

When the patient registries started, none of their founders planned to maintain the registries for only a short term (several years). One patient registry was scheduled to review the operational plan every two years. One patient registry was planned for a certain period of time (20 years). The remaining four were planned or sought to maintain for an unlimited period (Table 5). Of five registries that were planned to operate for a long time, three answered that they had no prospect of securing a budget in the future (Table 6).

**Table 5.** Planned operation period for operating patient registries at establishment

|  | Number of patient registries |
| --- | --- |
| A. No specific plan | 0 |
| B. Until research fund is finished | 0 |
| C. After a certain period | 1* |
| D. Permanently | 3 |
| E. Others | 2†,‡ |

\* Twenty years
† The project will be updated every two years.
‡ The research grant is for two years. I am not sure, but hope the registry will continue even after the grant period.

**Table 6.** Prospect of securing budget for long-term maintenance of patient registries when responding this survey

|  | Number of patient registries |
| --- | --- |
| A. Not planned long-term operation | 1 |
| B. Planned long-term operation, but with no prospect of achieving budget | 3 |
| C. Planned long-term operation, with prospect of achieving budget | 2 |

### Ideas and difficulties in building and sustaining patient registries

Finally, the questionnaire asked respondents to answer, by free description, questions related to 1) ingenuity and attention, 2) difficulty and anxiety, and 3) other free comments for building and maintaining each patient registry.

For ingenuity and attention, efforts of three kinds were raised from the respondents. The first type of answer was to endeavor to reduce burdens on registrants (medical doctors and patients) to register through keeping the number of follow-up surveys to once a year, including a reply envelope with survey form, or offering a toll-free phone number of the registration office. The second type was to achieve understanding of the importance of registration through organizing lecture meetings throughout the country, sending feedback and disclosing information to registrants and registry users such as researchers at public institutions or private companies, or carefully gathering feedback from them. The last type was to improve the number of registrations and persistence rates of registration through cooperation with patient groups and medical doctors throughout the country, or continually showing the passion of the head researcher of the registry by regularly sending letters or e-mails.

Regarding difficulty and anxiety, answers of five kinds were received. The first were related to securing a continuous budget. When the patient registry budget depends heavily on public grants from government ministries, the research group that manages the registry must apply for the grant every several years. It is therefore difficult for patient registries to establish stable and sustainable operation. The second were related to managing the burdens of the registration bureau. Continuing patient registry is currently dependent on the ambition and responsibility of the head researcher of the registry because of a lack of sufficient government support. Breaks in public research grants occurring every several years request a change of the head and members of the research group, which makes it difficult to continue the patient registry. The third is about inspiring the cooperation of medical doctors at small hospitals or clinics. For some rare diseases, patients visit neighboring hospitals unless facing serious matters, but most doctors in such small-scale hospitals have insufficient time and capacity to fill in registration forms for the patient. The fourth is related to use of the data in the registry.

Data in patient registries are expected to be used for the research and development of new drugs at private companies. That is a major aim of patient registries. However, it has not yet occurred for most patient registries. The last is about taking the balance between the amount of information and the number of registrants. More numbers and variety of items in the registration form give more information related to the disease and patients with the disease, although it gives lower enrollment numbers and renewal rates in registration because of increasing burden of filling in the registration form.

Regarding other free comments, some opinions were offered to indicate the future directions of patient registries. 1) Versatile web-based registration systems, on which many research groups can construct their patient registry, are necessary to reduce the burdens of constructing and maintaining patient registries. 2) There should be ways and means to use dormant patient registries that have been closed by budget shortages. 3) Target diseases, research into which the public resources are injected, should be emphasized. Many of them remain more or less unknown. 4) A trend shifting from a doctor-centered patient registry to a patient-centered one is expected to change the current undesirable circumstances related to the patient registry. 5) It would be better to try to introduce a new support scheme to achieve budgets for maintaining patient registries from pharmaceutical industry groups.

## Discussion

Results of this survey highlight current efforts and needs for developing and maintaining patient registries for rare diseases in Japan. This report is the first of a survey on this topic conducted in Japan. Although the respondents were few, qualitative tendencies were observed in terms of the results.

Some patient registries have no website. Therefore an internet search of patient registries is insufficient to find them, but the methodology used in this survey, using not only an internet search but also a database of grant-funded projects, is expected to address that limitation. The total number of rare disease patient registries in Japan is assumed to be small. Therefore, efforts for increasing the response rate will be important for future surveys.

The years of foundation of the patient registries to which the questionnaires were sent were distributed throughout 1997–2011, although those of the respondents were 2007–2011. It is not certain but probable that all or most patient registries established before 2007 were unable to sustain their activities beyond five years.

### Objectives of patient registries

Most respondents selected choices related to getting an overview and insight into diseases as objectives of the patient registries, although few selected ones related to ascertaining the real conditions of patients with diseases in their daily life, to assess the quality of medical care, or to research markets for drugs. This trend is the same as that in the case of Europe. In the EPIRARE survey [EPIRARE 2013], 70.8% of the respondent patient registries chose “epidemiological research” as the aim of the registry, although 33.3% chose “treatment monitoring” and 19.2% chose “social planning”.

Drugs and medical devices can improve the life experiences of patients. To achieve that improvement continuously, promotion of industry–government–academia cooperation would play an important role. The MHLW in Japan proposed “Clinical Innovation Network (CIN)” in 2015 as a new scheme for collaboration among six National Centers (NCs) for Advances and Specialized Medical Care affiliated with the MHLW and pharmaceutical companies. In this scheme, the NCs are presumed to construct patient registries as infrastructure to promote drug development [MHLW 2015]. The CIN would serve as a test of the patient registry for drug development.

### Structure and costs for operation

Half of the respondent patient registries chose “1,000,000–5,000,000 JPY” as the total expense for building and maintaining patient registries. The second most common answer was “5,000,000–10,000,000 JPY”. Expense items were budgets for labor, IT systems, and office supplies in descending order. In the EPIRARE survey, the most common answer to the question “Average yearly budget over three years” was “<50,000 EUR” (23.8% of respondents chose it). The third most common answer was “51,000–100,000 EUR” (13.4%). Fifty thousand EUR was about 51 million JPY in 2012, trends in yearly budgets were similar between Europe and Japan, except that the second most common answer was “No funds” (22.3%) in the EPIRARE survey.

Half of the respondent patient registries employ one or more full-time staff members. All employ part-time staff members. Although contents of work probably vary greatly according to each registry, some or most of them can be commonalized. If it were achieved, then patient registries could share staff members, which will engender reduction of the total cost.

Of the six respondents, two answered that their patient registries use web-based registration forms. One of them uses both the web-based and paper-based forms. Although the web-based registration form has several beneficial features, some registrants (patients or medical doctors) do not use the internet, some medical doctors are unable to connect the internet from their offices because of security policy, and the web-based registration form is expensive. It is necessary to reduce the cost of web-based forms to increase the adoption of web-based forms by, for example, sharing web-based systems among many patient registries.

### Future directions of rare disease patient registries

From free descriptions, important needs were extracted: obtaining funds, reducing operational burdens, gaining cooperation from participants (patients and medical doctors), promoting utilization of stored data, and so forth. These were similar to those reported from the EPIRARE survey, which elucidated frequent needs as financial support, strengthened motivation of data providers, data quality assessment, improvement of communication, and visibility of the results obtained. Here, we propose three cooperative efforts to address these issues.

First, a possible effort to reduce burdens related to constructing and maintaining patient registries is building a common platform that offers universal facilities such as IT infrastructure and secretariat functions. Many tasks are not specific to each patient registry. They might be outsourced or shared with many other registries. Such efforts would have a strong effect on expenses for constructing and maintaining patient registries because the IT infrastructure and labor cost are major expense items.

Second, patient registries should mutually collaborate in engaging in educational activities to increase awareness of the importance of patient registries. Public attitudes about participation in biobanks have a correlation with awareness of existence and the purpose of biobanks [Gaskell & Gottweis 2011]. Attitudes related to patient registries can show the same trend. Improved public awareness of patient registries is expected to increase the number of participants and the ratio of participants who engage continuously in patient registries and also increase the users of data stored in patient registries.

In Japan, a new law for medical care for patients with rare and intractable diseases was implemented on January 1, 2015. Based on this law, Japan’s MHLW has developed the rare and intractable diseases registry database to collect medical information of patients in Japan with rare diseases that are specified in this law. This registry is expected to improve the current circumstances. Data in this registry would be used for various purposes, although some researchers will continue to try to develop their own patient registries for purposes that are not accomplished with the MHLW’s registry and for diseases that are not included in the MHLW’s registry.

Third, a new scheme is expected to be introduced to secure a sufficient budget from collaboration between academia and industry to maintain patient registries. Data stored in patient registries is not necessarily useful and attractive for pharmaceutical companies and medical device companies because objectives to organize patient registries vary. However, founders of patient registries should devote more attention to supporting the development of new drugs and medical devices because they will have the potential to change a patient’s life drastically. Patient registries should consider collecting data that will attract researchers in academia but also those in industry at the planning phase in terms of increasing the use of data stored in registries and in terms of expanding the range of the sources of supporting funds.

As described previously, CIN was started in 2015 as a new collaboration framework between academia and industry to encourage drug discovery and development. It is drawing attention as a new framework for the use of patient registries.

## Conclusions

Survey results imply that costs and efforts to build and maintain patient registries for many rare diseases make them unrealistic. Some alternative strategy is necessary to reduce burdens, such as offering a platform that supplies IT infrastructure and basic secretariat functions that can be used commonly among many patient registries.

## Data Availability

na

## Abbreviations

BTMU: Bank of Tokyo-Mitsubishi UFJ
CIN: Clinical Innovation Network
EPIRARE: European Platform for Rare Diseases Registries
EUCERD: European Union Committee of Experts on Rare Diseases
EUR: Euro
GBP: Pound Sterling
IT: Information Technology
JPY: Japanese Yen
MHLW: Ministry of Health, Labour and Welfare
MURC: Mitsubishi UFJ Research and Consulting
NCs: National Centers
RDTF: Rare Diseases Task Force
TTB: Telegraphic Transfer Buying rate
TTS: Telegraphic Transfer Selling rate
USD: US Dollar

## Declarations

## Acknowledgments

The authors thank all responders of the survey for their contribution of the study. The authors are also grateful to Rie Momose for her valued support with survey conduct and data processing.

## Availability of data and materials

Not applicable.

## Authors’ contributions

MM conceived of the study and wrote the manuscript. MM and SO carried out the analysis. All authors read and approved the final manuscript.

## Ethics approval and consent to participate

Not applicable.

## Consent for publication

Not applicable.

